# Racial/Ethnic Heterogeneity in Diet of Low-income Adult Women in the United States: Results from National Health and Nutrition Examination Surveys 2011-2018

**DOI:** 10.1101/2022.04.06.22273539

**Authors:** Briana Joy K. Stephenson, Walter C. Willett

## Abstract

**Background:** Poor diet is a major risk factor of cardiovascular and chronic diseases, particularly for low-income women. However, the pathways by which race/ethnicity plays a role in this risk factor have not been fully explored.

**Objective:** This observational study aims to identify dietary consumption differences by race/ethnicity of US women living at or below the 130% poverty income level from 2011-2018.

**Design:** A total of 3005 adult women aged 20-80 years from the National Health and Nutrition Examination Survey (2011-2018) living at or below the 130% poverty-income level with at least one complete 24-hr dietary recall were classified into 5 self-identified racial/ethnic subgroups (Mexican, Other Hispanic, Non-Hispanic White, Non-Hispanic Black, Non-Hispanic Asian). Dietary consumption patterns were defined by 29 major food groups summarized from the Food Pattern Equivalents Database and derived via a robust profile clustering model which identifies foods that share consumption patterns across all low-income adult women, and foods that differ in consumption patterns based on race/ethnic subgroups.

**Results:** Legumes (protein and vegetable) were the most differentiating foods identified across all racial/ethnic subgroups and were primarily consumed by Mexican and Other Hispanic women. Non-Hispanic Asian women were most likely to favor a high consumption of prudent foods (fruits, vegetables, whole grains). Non-Hispanic White and Black women shared the most similarities in consumption patterns but differed in foods such as milk, poultry, and eggs.

**Conclusions:** Differences among consumption behaviors of low-income women were found along racial/ethnic lines. Efforts to improve nutritional health of low-income adult women should consider racial/ethnic differences in diet to appropriately focus interventions.

**Disclaimers:** N/A.

**Sources of Support:** Study supported in part by National Heart Lung and Blood Institute (NHLBI) grant R25 (HL105400) to Victor G. Davila-Roman and DC Rao.

## INTRODUCTION

Diet is one of the largest contributing factors to cardiovascular and other chronic diseases (Mokdad, 2018). The impact of cardiovascular disease (CVD) related outcomes on women aged 35-54 has increased in the past twenty years, while decreasing among other subgroups (Kuehn, 2019; Kalinowski, 2019; Thompson, 2017; Van Dyke, 2018). This gender disparity has been suggested to result from differences in diagnosis, care, and research specifically focused on women populations (Kuehn, 2019). CVD research that ignores gender differences create misleading extrapolation and a gap in understanding women-specific risk factors. Amidst this gender gap, racial and economic disparities have also continued to persist. Reportedly, 45% of Black women have some type of CVD, compared to 32% of white women (Williams, 2009). As disparities in CVD health continue to widen, the pathways through which factors like gender or race/ethnicity contribute to these differences are still not fully understood (Winkleby, 1999).

Dietary intake and behaviors has been widely studied in an attempt to identify modifiable pathways for improving overall health and preventing chronic diseases. Many studies have focused on dietary quality, using standardized adherence scores (e.g. Healthy Eating Index, Mediterranean Diet Score) (Hu, 2020; Onvani, 2017; Pate, 2015). While others have focused on individual foods/nutrients to examine diet-disease relationships (Basu, 2013; Drewnowski, 2013).

Foods and/or nutrients are commonly consumed together and share an intercorrelation structure, and biological synergy may exist among dietary components. For these reasons, dietary patterns can be a useful way to define exposure in studies of diet–disease relationships, complementing those of specific foods or nutrients (Hu, 2000; Schulze, 2018). Defining patterns by consumption of specific foods can also provide a simplified way to identify dietary behaviors of subgroups.

Latent class models are an easy and interpretable technique to identify dietary patterns, based on the consumption or nonconsumption of a large set of foods. However, this technique relies on a population composition where the single differentiating behavior present is diet. Populations that contain a mixture of demographics (e.g., culture, gender, SES) that may mediate differences in dietary behaviors pose challenges to the standard structure of latent class models. If a certain mediating demographic comprises the majority of the population, behaviors reflected in those dietary patterns will be dominated by the larger demographic. Smaller-sized demographics may have differing features, but are often overlooked and treated as noise. The focus of analysis is controlled by the identifiable behaviors present in the larger-sized demographic. For example, in many national surveys and cohorts of adults in the United States, the study population is often a majority of non-Hispanic white participants living above the 130% poverty income level. Modeling dietary behaviors from study populations sharing this makeup will yield behaviors reflective of this demographic, and prevent us from better understanding factors impacting the populations at greatest risk for health outcomes (racial/ethnic minority and low-income) (Stephenson, 2021; Field, 2007; Gavin, 2011)

More flexible model approaches have recently been introduced to improve the way we consider demographic features that may drive dietary behaviors. Robust Profile Clustering (RPC) is an extension to the latent class model that distinguishes consumption patterns that may be shared across the study population, and those that may be specific to a defined demographic. The technique has previously been applied to identify differences in maternal diet by geography, as well as adult diet of Hispanics/Latinos based on cultural background and US residency (Stephenson, 2019; Stephenson, 2020). With a focus on low-income adult women, this paper aims to apply the RPC model to identify racial/ethnic differences amongst an at-risk demographic.

## METHODS

### National Health and Nutrition Examination Survey

National Health and Nutrition Examination Survey (NHANES) is a publicly available population-based repeated cross-sectional survey. Approximately 9,000 people are sampled annually from 15 unique counties of varying socioeconomic and racial/ethnic backgrounds. Survey sampling weights are provided to generate population-based estimates of collected measures.

Our analysis focused on low-income female participants aged 20 and over who self-identified as Mexican-American, Other Hispanic, Non-Hispanic White, Non-Hispanic Black, Non-Hispanic Asian. Participants who identified as Mixed/Other or missing race/ethnicity information were excluded. Low-income was defined as those reporting a ratio of family income to poverty level at or below the 130%. We pooled four survey cycles for analysis (2011-2012, 2013-2014, 2015-2016, 2017-2018). Sampling weights were adjusted for pooled analysis in accordance with NHANES survey methods and guidelines (Johnson, 2014; Chen, 2018; Chen, 2018).

Dietary intake was measured using two 24-hour dietary recalls for each participant, collected initially in person, and the second recall was collected via telephone three to ten days later. Participants with at least one complete dietary recall were included for analysis.

Participants with two complete recalls available, were averaged over the two days. Recalls were collected as part of the What We Eat in America survey component of NHANES (Bodner-Montville, 2006). Nutrient composition for all foods and beverages reported were calculated using the Food and Nutrition Database for Dietary Studies (FNDDS). These nutrients were then converted into food pattern equivalents in accordance with the Dietary Guidelines for Americans (Bowman, 2014; Bowman, 2017; Bowman, 2018; Bowman, 2020). The food pattern equivalents database summarized the diet data into 29 major food groups. Levels of consumption for each food group were categorized into four levels, no consumption, low consumption (lower tertile of positive consumption), medium consumption (middle tertile of positive consumption), high consumption (upper tertile of positive consumption), where tertiles were calculated based on the overall population (Liu, 2019; Sotres-Alvarez, 2010).

CVD risk factors were treated as binary outcomes. High cholesterol was defined as participants with more than 200 total cholesterol or LDL cholesterol > 150 mg/dL or self-report taking cholesterol lowering medication. Obesity was defined as participants with a BMI > 30 kg/m^2^. Hypertension was defined as participants with an average blood pressure reading of systolic blood pressure greater than 140 mg/dL, diastolic blood pressure greater than 90 mg/dL, or self-reported medication use of blood pressure lowering medication. Diabetes is defined as participants with >126 mg/dL or self-reported diabetes medication use. Current smoker is defined as those who responded yes to the question “SMQ040: Do you now smoke cigarettes?”. Participants with complete diet data but missing one or more CVD risk factor data were still included for analysis but not included in the descriptive reporting for that missing risk factor.

### Robust Profile Clustering

Robust Profile Clustering (RPC) is a flexible extension of the latent class model. First introduced in Stephenson et al., the model breaks apart the assumption that all participants assigned to the same latent class (diet pattern) will share the same consumption behaviors for all food items observed (Stephenson, 2019). Alternatively, the RPC allows participants who share overall consumption patterns of a subset of foods to assume a global pattern, and participants who share unique consumption patterns within a predefined subgroup of the population assume a localized pattern.

### The model assumes a probabilistic framework comprised of three components

1. *Global pattern assignment*. This is consistent with the standard latent class model, where a global pattern describes the latent dietary behaviors shared amongst the overall population. The assignment of the global pattern is determined through a probability vector that describes the probability of a person being assigned to one global pattern over another.
2. *Local pattern assignment*. Within each subpopulation there exists another set of latent dietary behaviors that share attributes specific to their subpopulation, referred here as local patterns. The assignment of the local pattern is determined through a probability vector that describes the probability of a person from this subpopulation being assigned to one local pattern over another.
3. *Global/Local indicator*. Each food item used to describe each respective pattern has a probability of assuming the pattern detailed at the global level or the local level. This indicator, which is unique for each participant in the study population, is determined through a probability describing the likelihood of this food item to assume a global versus local pattern for that individual. The lower the probability the more likely it is to assume a local pattern. The higher the probability the more likely it is to assume a global pattern.

Each dietary pattern (global or local) is described with a probability matrix of *p* food item rows, where each row describes the probability of consumption at each of the *d* possible consumption levels. The modal pattern for each profile can be identified by the consumption level category with the highest probability value in that row.

We can describe the model mathematically, using the following notation. Let *y*_*i*_ = (*y*_*i*1_, …, *y*_*ip*_) denote the self-reported levels of consumption of *p* unique food/beverage items by individual *i* ∈ (1, …, *n*) where n is the total number of participants in the study population. Let 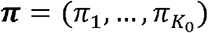 denote the probability vector for global pattern assignment, and 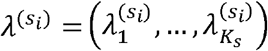 denote the probability vector of local pattern assignment where *S*_*i*_ ∈ (1, …, *S*) indicates the subpopulation index of individual *i* ∈ (1, …, *n*). Let *θ*_0*j* · |*h*_ *=* (*θ*_(0*j*1|*h)*,_ …, *θ*_(0*jd*|*h)*_) denote the consumption *d-*length probability vector for food item *j* ∈ (1, …, *p*) given assignment to global pattern *h*. Let 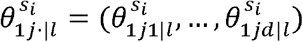 denote the consumption probability vector for food item *j* ∈ (1, …, *p*), given assignment to local pattern *l* within subpopulation index *S*_*i*_ ∈ (1, …, *S*). Let *G*_*ij*_ denote the global/local indicator variable for individual *i* ∈ (1, …, *n*) and food item *j* ∈ (1, …, *p*). Finally, let *K*_0_ and *K*_*S*_ denote the number of global and local patterns, respectively. The subject-specific likelihood of the RPC model can be described as

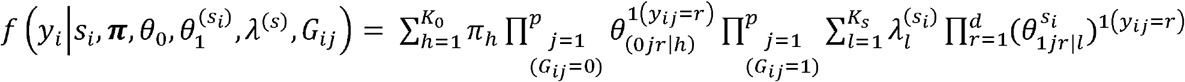

Estimation of the parameters of this model can be performed using a Bayesian approach. We fit the model using a Gibbs sampling algorithm, with conditional posterior distributions described previously (Stephenson, 2020). Noninformative, flat priors were selected to let the observed data drive model estimation. The number of global and local pattern were unknown *a priori*. As a result, we overfit the RPC model with 30 global and local patterns each so that when run using Markov chain monte carlo (MCMC), an interpretable set of nonempty global and local patterns would remain (Van Havre, 2015). Posterior computation, MCMC diagnostics, prior sensitivities, and convergence were performed as described previously (Stephenson, 2019; Stephenson, 2020). Data was preprocessed using SAS 9.4 (Cary Institute, 2013). RPC model was analyzed in Matlab 2021a. Survey sampling weights accounting for NHANES study design were incorporated *post hoc* and summarized using the survey package in R 4.0. All data and code to replicate analysis has been made publicly available on GitHub repository (https://github.com/bjks10/RPC/tree/master/NHANES).

## RESULTS

A total of 3005 female adult participants living at or below the 130% family poverty income level were included for analysis of this study (**Figure 1)**. Demographic information of these participants are provided in **Table 1**. Participants were mostly between the ages of 20-34 years of age (32.8%), self-identified as Non-Hispanic White (49.9%), married or living with a partner (45.9%), at least some college (43.6%). Most participants reported having high cholesterol.

**Figure 1.**
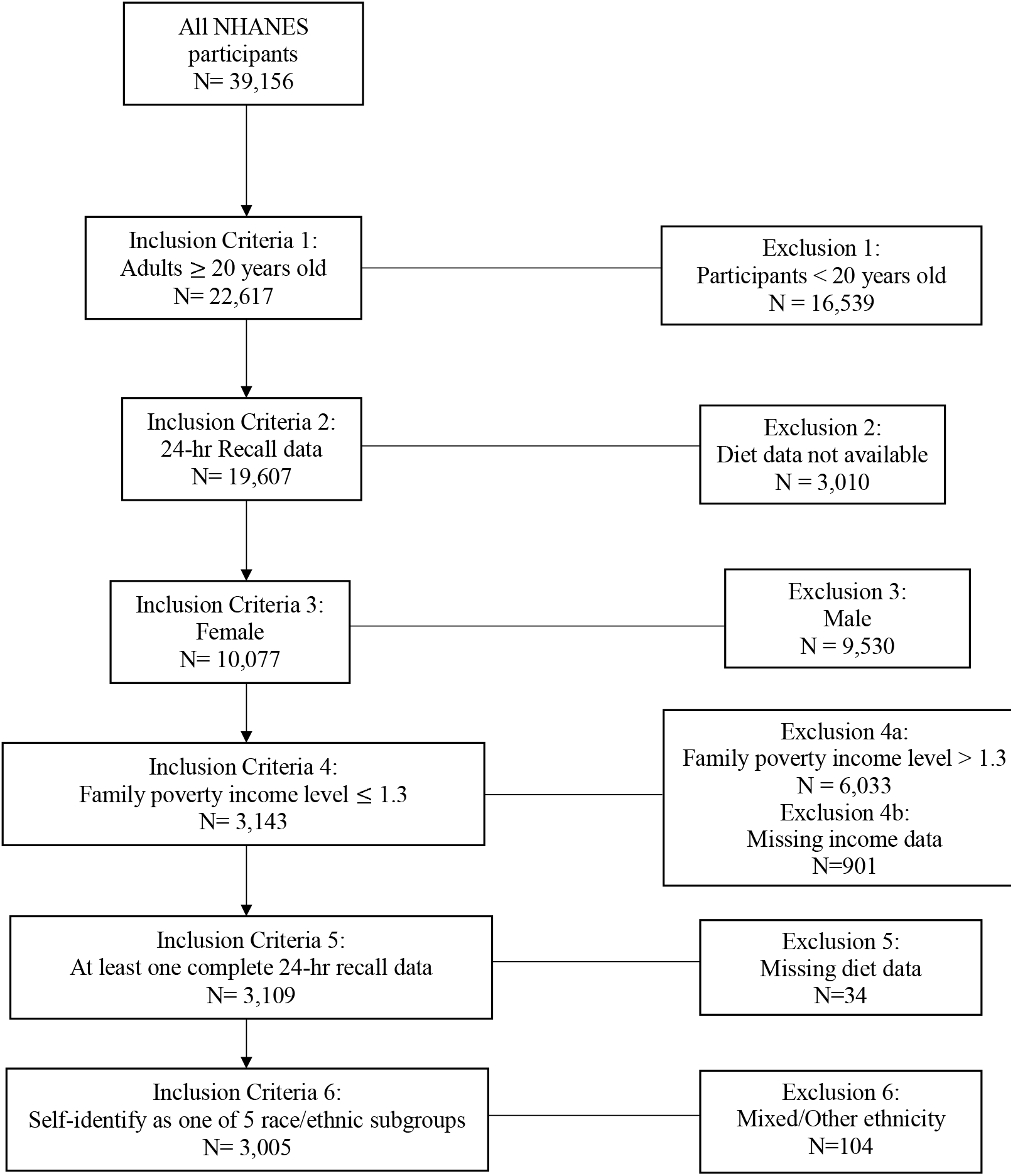
CONSORT diagram of NHANES 2011-2018 participants included for study analysis. A total of 3005 participants were included after all exclusion/inclusion criteria was applied.

**Table 1.**
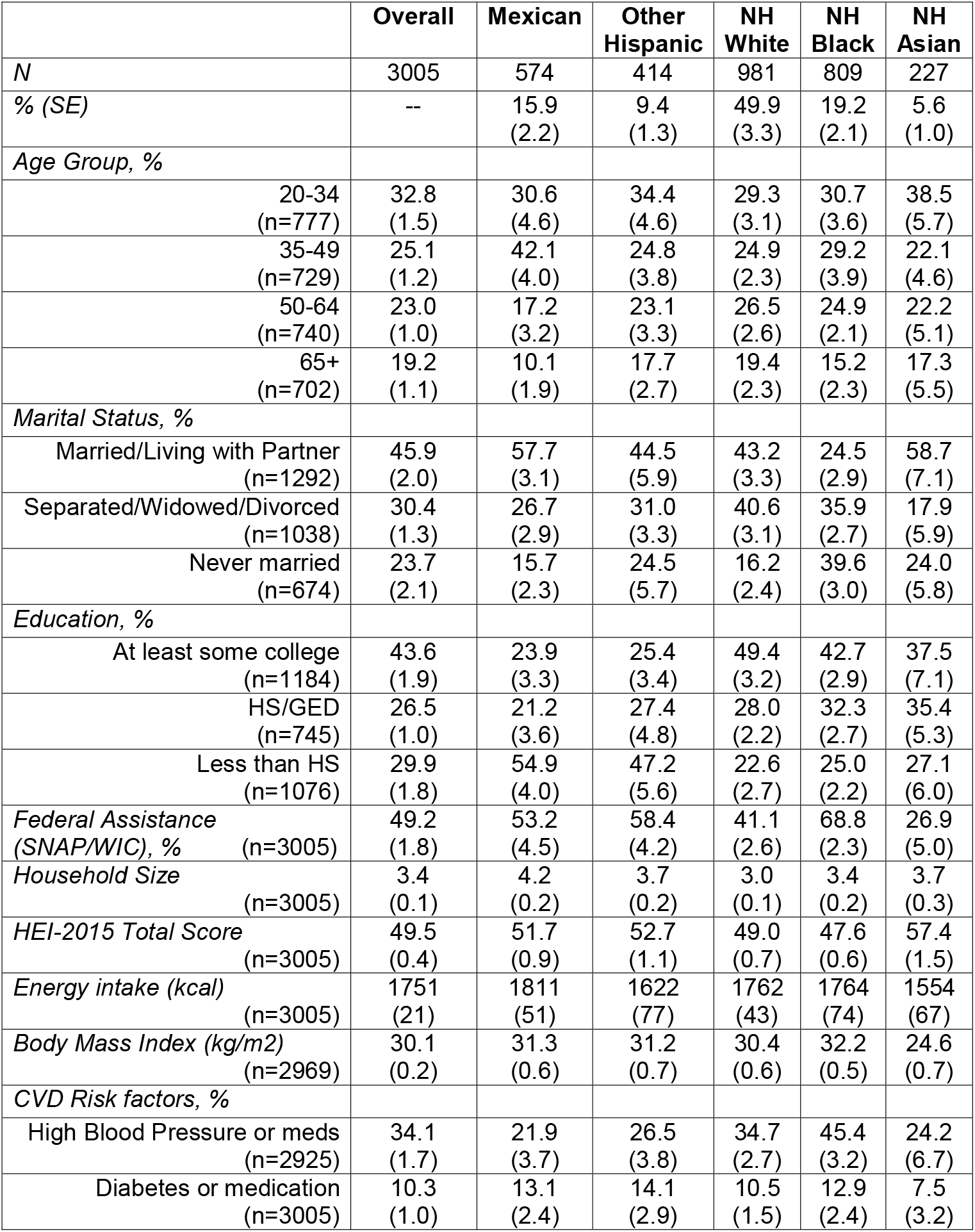
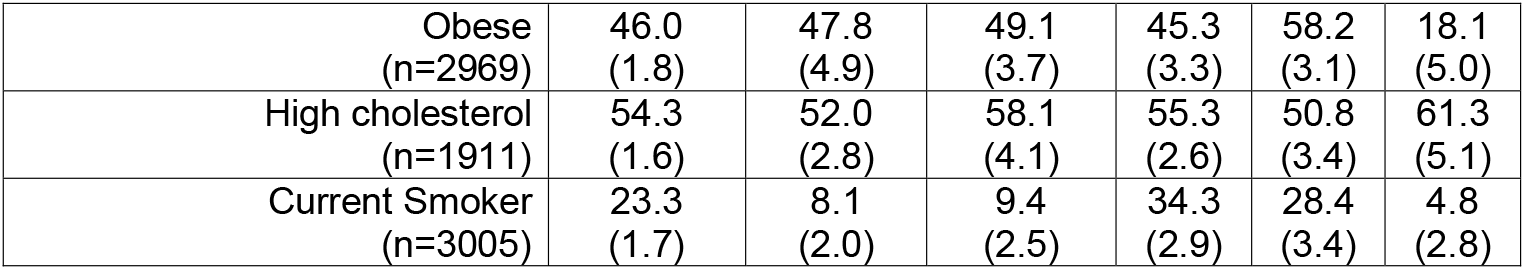
Demographic information of low-income adult women participants pooled from 2011-2018. Percentages account for survey sampling weights. Counts are unweighted.

|  | **Overall** | **Mexican** | **Other**<br>**Hispanic** | **NH**<br>**White** | **NH**<br>**Black** | **NH**<br>**Asian** |
| --- | --- | --- | --- | --- | --- | --- |
| *N* | 3005 | 574 | 414 | 981 | 809 | 227 |
| % (SE) | -- | 15.9<br>(2.2) | 9.4<br>(1.3) | 49.9<br>(3.3) | 19.2<br>(2.1) | 5.6<br>(1.0) |
| *Age Group, %* |  |  |  |  |  |  |
| 20-34<br>(n=777) | 32.8<br>(1.5) | 30.6<br>(4.6) | 34.4<br>(4.6) | 29.3<br>(3.1) | 30.7<br>(3.6) | 38.5<br>(5.7) |
| 35-49<br>(n=729) | 25.1<br>(1.2) | 42.1<br>(4.0) | 24.8<br>(3.8) | 24.9<br>(2.3) | 29.2<br>(3.9) | 22.1<br>(4.6) |
| 50-64<br>(n=740) | 23.0<br>(1.0) | 17.2<br>(3.2) | 23.1<br>(3.3) | 26.5<br>(2.6) | 24.9<br>(2.1) | 22.2<br>(5.1) |
| 65+<br>(n=702) | 19.2<br>(1.1) | 10.1<br>(1.9) | 17.7<br>(2.7) | 19.4<br>(2.3) | 15.2<br>(2.3) | 17.3<br>(5.5) |
| *Marital Status, %* |  |  |  |  |  |  |
| Married/Living with Partner<br>(n=1292) | 45.9<br>(2.0) | 57.7<br>(3.1) | 44.5<br>(5.9) | 43.2<br>(3.3) | 24.5<br>(2.9) | 58.7<br>(7.1) |
| Separated/Widowed/Divorced<br>(n=1038) | 30.4<br>(1.3) | 26.7<br>(2.9) | 31.0<br>(3.3) | 40.6<br>(3.1) | 35.9<br>(2.7) | 17.9<br>(5.9) |
| Never married<br>(n=674) | 23.7<br>(2.1) | 15.7<br>(2.3) | 24.5<br>(5.7) | 16.2<br>(2.4) | 39.6<br>(3.0) | 24.0<br>(5.8) |
| *Education, %* |  |  |  |  |  |  |
| At least some college<br>(n=1184) | 43.6<br>(1.9) | 23.9<br>(3.3) | 25.4<br>(3.4) | 49.4<br>(3.2) | 42.7<br>(2.9) | 37.5<br>(7.1) |
| HS/GED<br>(n=745) | 26.5<br>(1.0) | 21.2<br>(3.6) | 27.4<br>(4.8) | 28.0<br>(2.2) | 32.3<br>(2.7) | 35.4<br>(5.3) |
| Less than HS<br>(n=1076) | 29.9<br>(1.8) | 54.9<br>(4.0) | 47.2<br>(5.6) | 22.6<br>(2.7) | 25.0<br>(2.2) | 27.1<br>(6.0) |
| *Federal Assistance*<br>*(SNAP/WIC), %*<br>(n=3005) | 49.2<br>(1.8) | 53.2<br>(4.5) | 58.4<br>(4.2) | 41.1<br>(2.6) | 68.8<br>(2.3) | 26.9<br>(5.0) |
| *Household Size*<br>(n=3005) | 3.4<br>(0.1) | 4.2<br>(0.2) | 3.7<br>(0.2) | 3.0<br>(0.1) | 3.4<br>(0.2) | 3.7<br>(0.3) |
| *HEI-2015 Total Score*<br>(n=3005) | 49.5<br>(0.4) | 51.7<br>(0.9) | 52.7<br>(1.1) | 49.0<br>(0.7) | 47.6<br>(0.6) | 57.4<br>(1.5) |
| *Energy intake (kcal)*<br>(n=3005) | 1751<br>(21) | 1811<br>(51) | 1622<br>(77) | 1762<br>(43) | 1764<br>(74) | 1554<br>(67) |
| *Body Mass Index (kg/m2)*<br>(n=2969) | 30.1<br>(0.2) | 31.3<br>(0.6) | 31.2<br>(0.7) | 30.4<br>(0.6) | 32.2<br>(0.5) | 24.6<br>(0.7) |
| *CVD Risk factors, %* |  |  |  |  |  |  |
| High Blood Pressure or meds<br>(n=2925) | 34.1<br>(1.7) | 21.9<br>(3.7) | 26.5<br>(3.8) | 34.7<br>(2.7) | 45.4<br>(3.2) | 24.2<br>(6.7) |
| Diabetes or medication<br>(n=3005) | 10.3<br>(1.0) | 13.1<br>(2.4) | 14.1<br>(2.9) | 10.5<br>(1.5) | 12.9<br>(2.4) | 7.5<br>(3.2) |
| Obese<br>(n=2969) | 46.0<br>(1.8) | 47.8<br>(4.9) | 49.1<br>(3.7) | 45.3<br>(3.3) | 58.2<br>(3.1) | 18.1<br>(5.0) |
| High cholesterol<br>(n=1911) | 54.3<br>(1.6) | 52.0<br>(2.8) | 58.1<br>(4.1) | 55.3<br>(2.6) | 50.8<br>(3.4) | 61.3<br>(5.1) |
| Current Smoker<br>(n=3005) | 23.3<br>(1.7) | 8.1<br>(2.0) | 9.4<br>(2.5) | 34.3<br>(2.9) | 28.4<br>(3.4) | 4.8<br>(2.8) |

Local patterns were defined by five racial/ethnic subgroups. The demographic characteristics of these racial/ethnic subgroups are described in Table 1. Non-Hispanic Black women were more likely to be never married, whereas all other racial/ethnic subgroups were more likely to be married or living with a partner. Mexican and Other Hispanic women were more likely to have less than a high school education. The other three racial/ethnic subgroups were more likely to have at least some college education.

### Global Dietary Profiles

Five global patterns were derived. **Figure 2** illustrates the modal pattern of consumption for each profile. Global profiles 1 and 2 have the most similarities, sharing consumption modes for 22 of 29 food items. Both profile patterns favor a high intake of cheese, oils, fats, and sugars, as well as grains, other vegetables, and legumes. They favor no consumption of potatoes, other starchy vegetables, soybean products and alcohol. Where they differ is in the consumptions of meats and fruits. Global profile 1 favored a higher consumption of fruits, juices, high n-3 seafood, poultry, and green vegetables. Global profile 2 favored a higher consumption of organ meat and other red/orange vegetables. Global profiles 3 and 4 shared 14 consumption modes. These two profile patterns both favored a medium intake of other fruits, dark green vegetables, soybean products, oils, and sugars. However, global profile 3 favored a higher consumption of tomatoes, refined grains, eggs, and cheese. Global profile 4 comparatively favored a high consumption of organ meat. Global profile 5 had the least diet diversity favoring consumption of only 7 foods. Of these, poultry and other starchy vegetables favored a high level of consumption, while the remaining five foods (refined grains, soybean, oils, solid fats, and added sugar) favored a low level of consumption.

**Figure 2.**
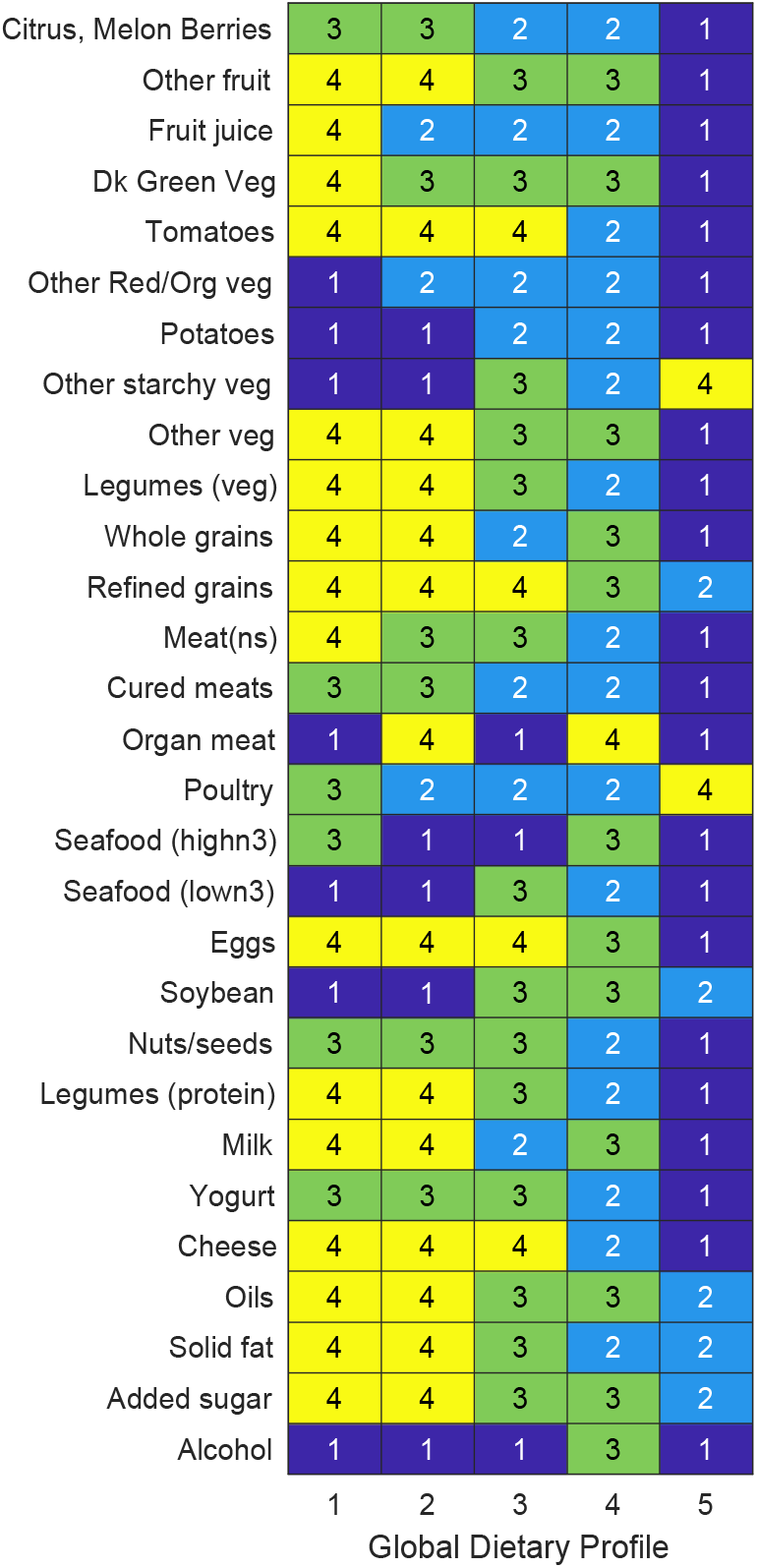
Consumption modal pattern for five RPC-derived global dietary profiles. Levels of consumption are highlighted as follows: 1-Dark Blue (no consumption), 2-Light Blue (low consumption), 3-Green (medium consumption), 4-yellow (high consumption). Key features of each global dietary profile: Global 1, high legumes, medium poultry; Global 2, high legumes, high organ meat; Global 3, medium legumes, medium seafood; Global 4, high organ meat, low fruit/veg; Global 5, high starchy vegetable, high poultry.

The distribution of these global patterns amongst the study population is provided in **Table 2**. Of immediate note is that global profile 5 contains about 70 percent of the population. This is likely due to the more populous racial/ethnic subgroups, Non-Hispanic White and Black women being assigned to this group. Mexican women were more likely to be assigned to global profile 2. Other Hispanic women were more likely to be assigned to global profile 3. Non-Hispanic Asian women were more likely to be assigned to global profile 4. Women in global profile 1 were more likely to be married or living with a partner and have less than a high school degree and had the largest average household size. Women assigned to global profile 2 averaged the highest healthy eating index score, whereas those assigned to global profile 5 averaged the lowest.

**Table 2.** Weighted means with standard errors in parentheses are provided to describe demographic distribution across RPC-derived global profiles.

|  | Global 1<br>8.9 (0.7) | Global 2<br>2.2 (0.4) | Global 3<br>8.9 (0.8) | Global 4<br>9.4 (0.7) | Global 5<br>70.5 (1.2) |
| --- | --- | --- | --- | --- | --- |
| *Age Group* |  |  |  |  |  |
| 20-34 | 33.1 (4.0) | 24.4 (6.9) | 33.0 (4.7) | 37.0 (4.3) | 32.4 (1.7) |
| 35-49 | 34.3 (4.6) | 40.9 (9.5) | 28.5 (4.2) | 20.2 (3.0) | 23.6 (1.3) |
| 50-64 | 19.3 (3.1) | 25.0 (6.7) | 22.0 (2.9) | 19.6 (3.0) | 24.0 (1.3) |
| 65+ | 13.4 (2.3) | 9.6 (3.7) | 16.5 (3.6) | 23.2 (3.4) | 20.0 (1.5) |
| *Race/Ethnicity* |  |  |  |  |  |
| Mexican | 29.5 (5.5) | 31.7 (7.5) | 23.8 (5.9) | 19.0 (5.2) | 11.2 (2.1) |
| Other Hispanic | 15.2 (4.5) | 3.0 (1.7) | 23.6 (6.6) | 10.6 (4.0) | 7.2 (1.3) |
| Non-Hispanic White | 43.7 (7.0) | 50.6 (11.1) | 40.2 (7.4) | 50.9 (7.4) | 55.5 (4.2) |
| Non-Hispanic Black | 7.7 (2.9) | 7.8 (4.4) | 9.5 (3.4) | 11.2 (3.0) | 21.1 (2.7) |
| Non-Hispanic Asian | 4.0 (1.6) | 6.9 (4.7) | 2.9 (1.3) | 8.3 (4.1) | 5.0 (1.1) |
| *Marital Status* |  |  |  |  |  |
| Married/Living with Partner | 67.8 (5.5) | 26.1 (9.1) | 45.9 (6.1) | 47.1 (5.9) | 38.3 (2.6) |
| Separated/Divorced/Widowed | 17.5 (4.7) | 51.3 (11.0) | 32.0 (5.8) | 32.3 (4.9) | 37.4 (2.1) |
| Never married | 14.7 (4.5) | 22.5 (9.2) | 22.0 (6.7) | 20.6 (5.3) | 24.4 (2.5) |
| *Education* |  |  |  |  |  |
| At least some college | 39.5 (5.8) | 52.2 (9.9) | 33.8 (4.9) | 45.4 (6.9) | 43.1 (2.4) |
| High School/GED | 17.9 (4.5) | 19.9 (7.9) | 27.9 (4.5) | 30.6 (6.4) | 29.1 (1.7) |
| Less than High school | 42.6 (5.9) | 27.9 (8.2) | 38.3 (5.7) | 23.9 (5.0) | 27.9 (2.0) |
| Federal Assistance Program (SNAP/WIC) | 53.2 (5.1) | 38.6 (9.3) | 58.5 (4.1) | 43.4 (4.2) | 48.6 (1.8) |
| *Continuous measures* |  |  |  |  |  |
| Household size | 4.0 (0.3) | 3.4 (0.4) | 3.8 (0.2) | 3.3 (0.3) | 3.2 (0.1) |
| HEI2015 Total Score | 55.3 (1.1) | 59.6 (1.5) | 51.2 (1.2) | 52.6 (1.4) | 48.2 (0.5) |
| Energy (kcal) | 2144 (74) | 2075 (142) | 1831 (87) | 1738 (68) | 1680 (31) |
| Body Mass index | 30.5 (0.7) | 33.5 (2.5) | 29.5 (1.1) | 28.1 (1.0) | 31.0 (0.4) |
| *CVD risk factors* |  |  |  |  |  |
| High blood pressure + meds | 27.8 (5.2) | 33.4 (9.5) | 30.2 (4.3) | 33.7 (5.0) | 33.2 (2.2) |
| Diabetes + meds | 12.0 (4.2) | 19.9 (7.5) | 6.2 (1.9) | 10.8 (3.5) | 11.6 (1.2) |
| Obese (BMI > 30) | 44.0 (5.9) | 48.4 (9.5) | 41.8 (5.2) | 30.3 (6.3) | 49.3 (2.2) |
| High Cholesterol | 76.6 (4.1) | 77.5 (7.1) | 69.8 (5.5) | 68.5 (6.3) | 72.5 (1.8) |
| Current smoker | 16.3 (5.0) | 26.6 (13.0) | 16.3 (4.2) | 18.7 (6.0) | 28.9 (2.2) |

### Racial/Ethnic-specific Local Patterns

As mentioned before, under the RPC model, women assigned to the global patterns are not beholden to all the consumption patterns described at the global level. To determine which food items assume which pattern, we must consider **Figure 3**. This figure provides a probability heatmap of a food item (y-axis) assuming the global pattern for each race/ethnicity subgroup (x-axis). Two food items had a higher probability for assuming a global pattern, legumes (protein) and legumes (vegetable). Mexican and Other Hispanic women had over 94% probability of assuming a global pattern. Recalling that the Mexican and Other Hispanic groups were more likely to be assigned to global profiles 2 and 3 we note that the mode of consumption for legumes in these profiles was a high and medium consumption level, respectively. Similarly, Non-Hispanic Asian women had a 74% probability of assuming the global patterns. These women had the largest representation in global profile 4, which had a high proportion of a low consumption of these food items. Both Non-Hispanic White and Black women were split in whether they would assume the global or local pattern. Focusing on their most representative global profile 5 consumption mode, and that for each of these subgroups at the local level, we see that they were most likely to not consume these food items at all.

**Figure 3.**
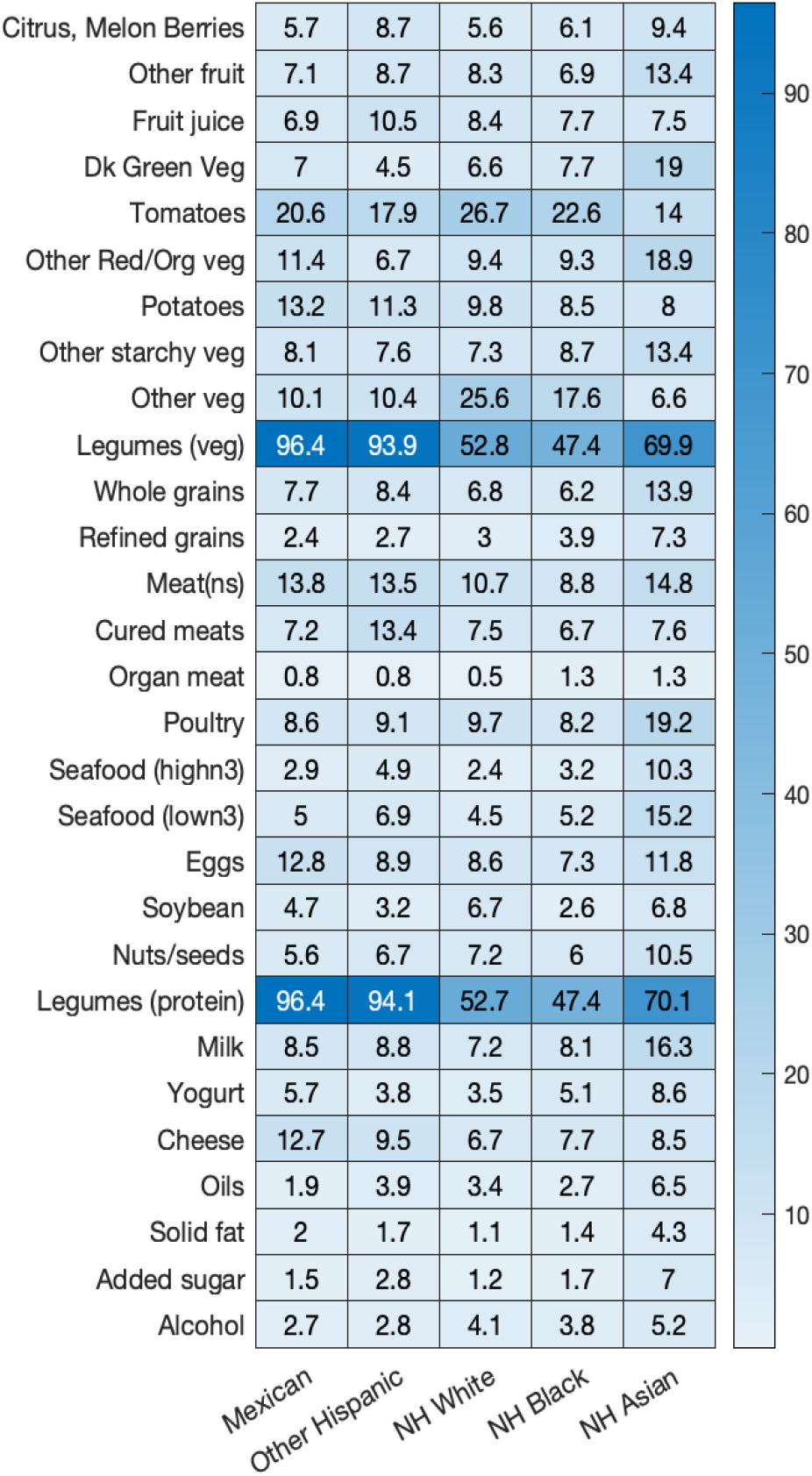
Heatmap illustrating probability of a given food item assuming a pattern at the global level. Foods likely to assume a global pattern are darker in blue hue. Foods likely to assume a localized pattern by the racial/ethnic subgroup are lighter in blue hue.

Given that the remainder of the food items were most strongly associated with a local pattern, we examine the locally stratified patterns for each of these racial/ethnic subgroups. **Figure 4** provides a full distribution plot of the pattern of each of the observed foods given assignment to each local profile. Each of the racial/ethnic subgroups had a single local profile to explain the different probabilities of consumption. Consumption modes are identifiable by the vertical bar of relative height.

**Figure 4.**
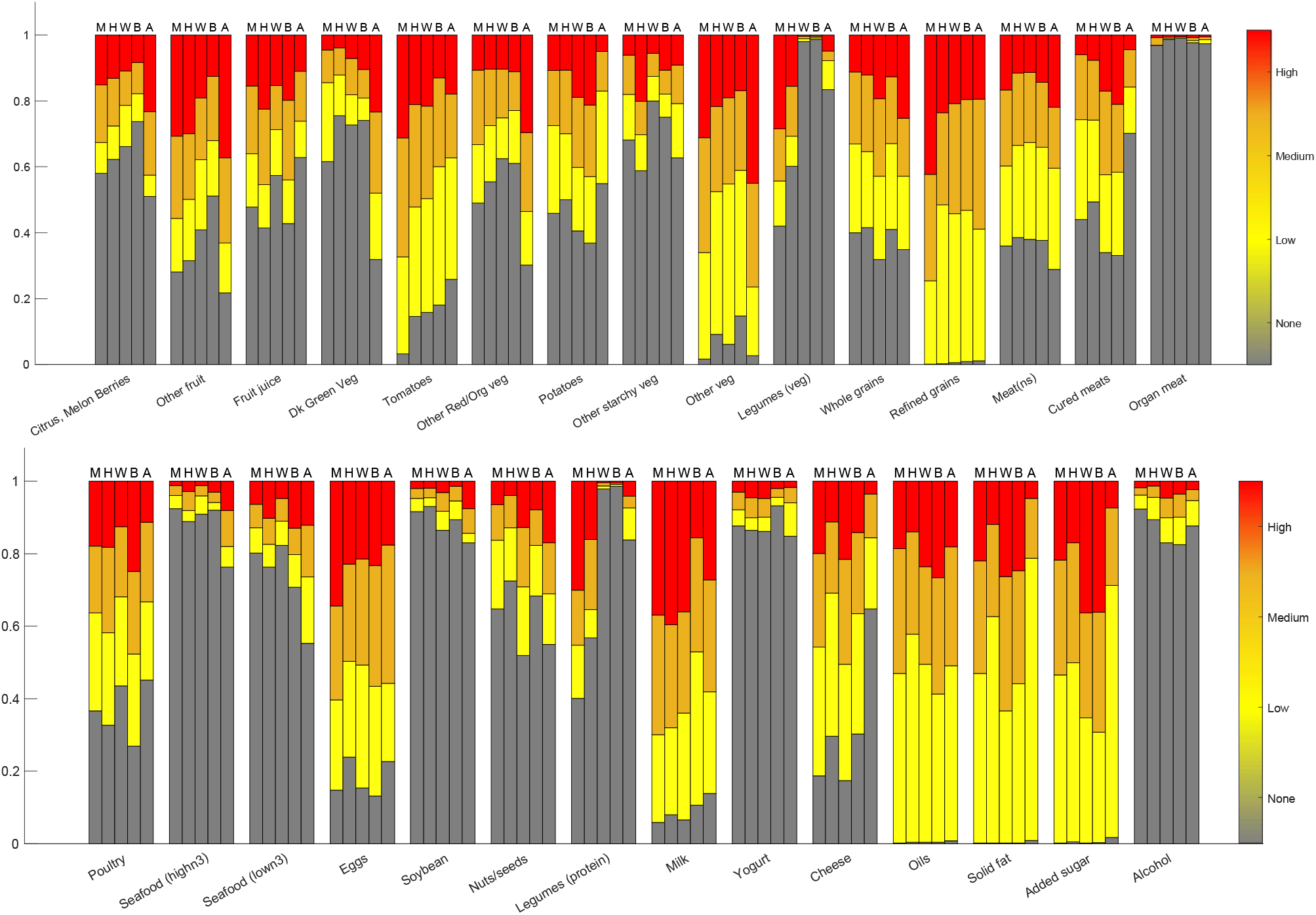
Pattern distributions for RPC-derived local profiles for each racial/ethnic subgroups. M=Mexican, H=Other Hispanic, W=Non-Hispanic White, B=Non-Hispanic Black, A=Non-Hispanic Asian.

Added sugars, solid fats, refined grains, and oils were consumed by all racial/ethnic subgroups. However, Non-Hispanic Asian women were the only subgroup that had some women who did not consume these foods at all during the two recall days of record. Mexican women were more likely to favor a high consumption of other fruit, refined grains, eggs, and milk, with a medium consumption of other vegetables and tomatoes. Other Hispanic women showed similar patterns to Mexican women, but favored a lower consumption of tomatoes, other vegetables, refined grains, and eggs. Non-Hispanic Black and Non-Hispanic White women showed the most similarities in their patterns. These two subgroups favored a low consumption of tomatoes, other vegetables, refined grains, cheese, and oils. Comparatively, Non-Hispanic Black women favored a high consumption of eggs (medium vs low), whereas Non-Hispanic White women favored a higher consumption of milk (high vs low) and solid fats (medium vs low). Non-Hispanic Asian had the most unique dietary pattern compared to the other four, with a high consumption of other fruit, other vegetables. While meats appear to have a mode of non-consumption for all groups, we see that Non-Hispanic Asian women were least likely to consume cured meats across subgroups, and most likely to consume non-specified meat and seafood. At the high consumption level, poultry and seafood (low-n3) were most likely to be consumed by Non-Hispanic Black women.

These local dietary patterns are consistent with the healthy eating index scores provided in Table 1. Average energy intake was highest amongst Mexican women and lowest amongst non-Hispanic Asian women. Non-Hispanic Asian women, whom had the highest probability of high consumption of foods like whole grains, other red/orange vegetables, and no consumption of cheese, milk, oils, solid fats, and added sugars, had the highest HEI score (57.4 +/- 1.5), and the lowest proportion of all five CVD risk factors. Both Non-Hispanic White and Black women had HEI scores lower than 50 with favored high consumption of starchy vegetables, refined grains, oils, and added sugars, and shared the highest proportion of women with high cholesterol. Non-Hispanic Black women also had the highest proportion of women with hypertension and obesity. Diabetes was most prevalent among Other Hispanic women.

## DISCUSSION

The application of this model allowed a deeper look into the consumption behaviors of adult women in the United States that live at or below the 130% poverty income level, and how those behaviors may differ by race/ethnicity. The standard latent class model relies on a homogeneous population where consumption habits may differ. Non-Hispanic White women are the larger racial/ethnic demographic. Consequently, this demographic and their consumption behaviors overpower the patterns reflected across all five global diet patterns. The RPC generated a more accurate identification of racial/ethnic dietary differences amongst these women.

Legumes (protein and vegetable) were the two most differentiating food features that were reflected across the five global patterns. Consumption of legumes (beans) has been associated with a myriad of nutritional benefits on lowering cardiovascular disease risk (Winham, 2009; Mattei, 2014; Winham, 2007; Finley, 2007; Bazzano, 2011; Thompson, 2012; Mitchell, 2009). However, the consumption patterns of these foods, continue to fall along cultural lines. Mexican and Other Hispanic women, where beans are a cultural staple of the home, were likely to have a high and medium consumption of legumes (Global patterns 1-3), respectively. Non-Hispanic Asian women, who had a higher representation in global pattern 4, were likely to have a low consumption. Non-Hispanic Black and White women, who made up the largest representation of the participants assigned to global pattern 5, were likely to not consume legumes at all.

Locally, we were able to see the consumption distribution of the remaining 27 food items by race/ethnicity. Unhealthy food items (refined grains, solid fats, added sugars) shared some level of consumption across all racial/ethnic subgroups, except for Non-Hispanic Asian women. Despite poorer expected outcomes and risk factors expected amongst low-income adults, Non-Hispanic Asian women had the healthiest of outcomes and lower prevalence of all CVD risk factors except cholesterol. The diets reflected in this subgroup had a higher proportion of high consumers of fruits, vegetables, and whole grains. Mexican and Other Hispanic women shared several similarities in consumption patterns across most foods, with a higher consumption by Mexican women of refined grains, tomatoes, eggs, and cheese. Non-Hispanic White and Black women also shared several similarities in consumption patterns, with higher consumption of poultry and eggs by Non-Hispanic Black women and lower consumption of dairy (milk, cheese).

Previous studies who examined the diets of low-income populations, have focused analysis on adherence scores to examine diet quality, emphasizing consumption of fruits, vegetables, and whole grains (Lin, 2005; Leung, 2012). Adjustments for intake or survey weighted means are adjusted using regression analysis focusing on specific foods or nutrients on independent models. Our analysis utilized the RPC model to examine the combination of all foods summarized by the dietary intake tool, under a latent class approach. Additionally, findings of prior studies indicated an overall lower intake of fruits and vegetables, whereas our joint-stratified model was able to identify certain racial/ethnic groups where healthier eating habits are present.

This analysis highlights one of the methodological strengths of the RPC model: 1) the ability to reduce the number of models to better understand the heterogeneity; 2) generating a joint-stratified latent class structure with a global LCA identifying patterns shared across multiple subgroups, and a stratified LCA to capture local patterns for each subgroup; 3) implementing an overfitted latent class model to determine the appropriate number of diet patterns, globally and locally in one single model.

With expansion of NHANES 2011 data collection, starting in 2011, our analysis was able to leverage additional racial/ethnic details (e.g. Non-Hispanic Asian, Mexican vs Other Hispanic) not previously available in prior NHANES diet research. This allowed a more in-depth analysis of dietary patterns shared across all low-income women, and across multiple racial/ethnic subgroups.

As with most models, the output is only as good as the input. Dietary patterns reflected in this model are based on the consumption amounts reported on two 24-hour dietary recalls by NHANES participants. Measurement error was not accounted for in this model, so cases of underreporting and overreporting are possible and patterns should be considered with potential misclassification in mind. As a result, the associations between race/ethnicity and diet patterns reported are likely to have been understated.

Food choices made by low-income women are often influenced by cost (Wiig, 2009). Federal assistance programs, such as Supplemental Nutrition Assistance Program (SNAP) and special SNAP for Women, Infants, and Children (WIC), have been created to assist in providing affordable options, but the selection of foods purchased and consumed may not be motivated with nutritional health in mind (Wardle, 2000). Wang et al. examined diet quality of adults from NHANES 1999-2010 and found that while income disparities accounted for differences in Non-Hispanic Black and White populations, it did not for other racial/ethnic subgroups (e.g. Mexican-Americans) (Wang, 2014). In other racial/ethnic subgroups, cultural influence can play an even more significant role in both diet and participation in federal assistance programs. For example, our study found that Non-Hispanic Asian women had the highest adherence to healthy habits and the lowest participation in SNAP or WIC federal assistance programs. The reasons behind these dietary differences and choices remain complex and open for further research.

This approach effectively identified heterogeneity in diet quality and foods consumed among subgroups of low-income women, defined by race/ethnicity, that have previously not been fully appreciated in overall population diet analysis. With a more recent focus on how to improve nutrition amongst federal assistance programs for low-income populations, approaches such as the RPC, may be of value for targeting efforts to improve diet quality within these subgroups and for a better understanding of health disparities overall.

## Data Availability

All data and supporting code to reproduce analysis are publicly available on GitHub repository: https://github.com/bjks10/RPC/tree/master/NHANES.

https://wwwn.cdc.gov/nchs/nhanes/default.aspx

https://www.ars.usda.gov/northeast-area/beltsville-md-bhnrc/beltsville-human-nutrition-research-center/food-surveys-research-group/docs/fped-databases/

## ACKNOWLEDGEMENTS

The authors thank the study participants of NHANES for their important contributions. The authors responsibilities were as follows: BJKS designed the research, analyzed, interpreted the data, wrote the manuscript, and had primary responsibility of content. WCW assisted with data interpretation, and critical revisions of the manuscript. All authors read and approved the final manuscript. This study was supported in part by NHLBI grant R25 HL105400 to DC Rao and Victor G. Davila-Roman.

## Abbreviations

CVD: cardiovascular disease
SES: socioeconomic status
RPC: robust profile clustering
NHANES: National Health and Nutrition Examination Survey
FNDDS: Food and Nutrition Database for Dietary Studies
LDL: low density lipoprotein
BMI: body mass index
MCMC: Markov Chain Monte Carlo
LCA: latent class analysis
SNAP: Supplemental Nutrition Assistance Program
WIC: women, infants, and children

